# On Gaps of Clinical Diagnosis of Dementia Subtypes: A Study of Alzheimer’s Disease and Lewy Body Disease

**DOI:** 10.1101/2021.05.05.21256720

**Authors:** Hui Wei, Arjun V Masurkar, Narges Razavian

**Author notes:** Corresponding author: Narges Razavian, 227 East 30th street Ste 639, New York NY 10016.

## Abstract

**BACKGROUND:** Alzheimer’s disease (AD) and Lewy body disease (LBD) are the two most common neurodegenerative dementias, and can occur in combination (AD+LBD). Due to overlapping biomarkers and symptoms, especially at early stages, clinical differentiation of these subtypes could be difficult. Yet it is unclear how the magnitude of diagnostic uncertainty varies across the dementia spectrum and demographic variables. We aimed to compare clinical diagnosis and post-mortem autopsy-confirmed pathological results to assess the clinical subtype diagnosis quality across these various factors.

**METHODS:** We studied data from the National Alzheimer’s Coordinating Center. Selection criteria included an autopsy-based neuropathological assessment for AD and/or LBD, and initial visit with Clinical Dementia Rating CDR® Dementia Staging Instrument (CDR) of 0, 0.5, or 1.0 corresponding to normal, mild cognitive impairment, or mild dementia, respectively. Longitudinally, we analyzed first visits at each subsequent CDR stage, and focused on the clinical diagnosis for AD and/or LBD in these visits. This analysis included positive predictive values (PPV), specificity and sensitivity and false negative rates of the clinical diagnosis, as well as disparities in these metrics by sex, race/ethnicity, age and education. If autopsy-confirmed AD, LBD, or AD+LBD was missed, the alternative clinical diagnosis was analyzed.

**FINDINGS:** Records of 1887 qualified participants from September 1, 2005 to August 20, 2019 were included in the study. Clinical diagnosis of AD+LBD had poor sensitivity from 2.7% (95% CI: 1.2%, 4.8%) at CDR 0.5 to 4.7% (2.1%, 7.8%) at CDR 3.0. Over 60% of participants with autopsy-confirmed AD+LBD were diagnosed clinically as AD. Clinical diagnosis of AD had low sensitivities at early dementia stage (56.8% (95% CI: 52.4%, 61.1%)) and low specificities at all stages. Among participants diagnosed as AD in the clinic, over 33% had concurrent LBD neuropathology at autopsy. Clinical diagnosis of LBD had poor PPVs from mild cognitive impairment (22% (95% CI: 12%, 33%)) to severe dementia (20% (95% CI: 9%, 33%)). Among participants diagnosed as LBD in the clinic, 32% to 54% revealed AD pathology during autopsy. When the three subtypes were missed by clinicians, “No cognitive impairment”, “Undecided” and “primary progressive aphasia or behavioral variant frontotemporal dementia” were the leading primary etiologic clinical diagnosis. With increasing dementia stages, the difference in clinical diagnosis accuracy significantly increased between White and Black/African-American participants, and diagnosis quality significantly improved for males but not for females.

**INTERPRETATION:** Our findings demonstrate that clinical diagnosis of AD, LBD and AD+LBD are inaccurate and harbor significant disparities based on race and sex. These findings have important implications for clinical management, anticipatory guidance, trial enrollment, and applicability of potential disease modifying therapies for AD. They also promote research into better biomarker-based assessment of LBD pathology.

## 1. Introduction

Alzheimer’s disease (AD) is the most common cause of dementia, accounting for 60% to 80% of dementia cases (Scheltens et al. 2016) (Soria Lopez, González, and Léger 2019). AD is neuropathologically defined by amyloid plaque and neurofibrillary tau tangles, and typically presents clinically as a progressive amnestic syndrome with minimal physical symptoms, especially in the early stages. Lewy body disease (LBD) is the second most prevalent neurodegenerative dementia subtype (Walker et al. 2015) (Mueller et al. 2017). In contrast to AD, LBD is neuropathologically linked to alpha synuclein-based Lewy body inclusions. It is clinically hallmarked by cognitive fluctuations, early onset visual hallucinations, REM sleep behavior disorder, and parkinsonism, with supportive features including atypical response to neuroleptics, autonomic dysfunction, and dysosmia (Ian G. McKeith et al. 2017).

Yet despite these seemingly disparate classical descriptions, in clinical practice there can be significant overlap between their presentations. AD and LBD can have similar neuropsychological profiles, especially considering dysexecutive and other variants of AD. Hallucinations tend to occur later in AD, but can sometimes appear at early stages. Parkinsonism can develop in the late stages of AD as well. Dysosmia is found in both AD and LBD. Lastly, APOE4 has been found to influence pure LBD pathology independent of AD pathology (Dickson et al. 2018). Moreover, AD and LBD can be found together on autopsy. This mixed subtype (AD+LBD) can lead to a more severe presentation (Förstl 1999) (Hansen 1997) with symptoms of both manifesting concurrently. Available clinical diagnostic criteria for AD (McKhann, Knopman, and Chertkow 2011) and LBD (I. G. McKeith et al. 2005) perform poorly when applied to this mixed dementia. How to identify and treat this mixed subtype is still an open research area, especially as there are biomarker-based assessments for AD but not LBD.

Earlier accurate diagnosis for dementia subtypes is critical for clinical care. Symptom management will be influenced by subtype, for example, leading to more caution with neuroleptic use in LBD or AD+LBD patients. For the benefit of the patient and their families, the anticipatory guidance for each subtype will differ as well. In the advent of disease-modifying therapies, especially in the case of AD, knowing the likelihood of concurrent LBD pathology will be important in determining whether such medications would be of value. This same consideration is applicable to ongoing and future clinical trials focusing on AD and LBD.

A number of previous studies assessed clinical diagnostic quality for dementia subtypes. (Beach et al. 2012) examined the clinical AD diagnosis accuracy of 919 participants at their last assessment during life from the 2005-2010 National Alzhimer’s Coordinating Center dataset. They concluded the clinical diagnosis is more accurate for AD than non-AD dementia. (Soria et al. 2018) found the same result for 53 Latino participants in UCSD Alzheimer’s Disease Research Center dataset from 1991 to 2017. Similarly, (Selvackadunco et al. 2019) measured the mismatch rate between clinical diagnosis and post-mortem neuropathological results of 7 dementia subtypes for 180 participants in Brains for Dementia Research cohort. They found the clinical misdiagnosis rate is high among all 7 subtypes. (Gianattasio et al. 2019) and (Lin et al. 2020) evaluated the racial dementia diagnosis disparity and found that Non-Hispanic Black participants have a higher underdiagnosis rate compared to Non-Hispanic White participants. Yet, these studies did not differentiate dementia stages or only focused on later stages. These results motivate a deeper analysis of diagnostic quality and disparity at all dementia stages separately to assess clinical diagnosis quality at earlier stages and whether it improves at higher stages.

In this paper, we focus on autopsy-confirmed diagnosis of 1887 dementia participants, and assess the clinical diagnosis made for them at different stages of their cognitive impairment (CDR 0.5, 1.0, 2.0 and 3.0). We focus on the sensitivity, specificity, and positive predictive value of AD, LBD, AD+LBD subtypes, as well as disparity measures. Our results demonstrate that the current clinical diagnosis accuracy is still low at all cognitive impairment stages for all dementia subtypes, and there are significant disparities based on race and sex. These findings have important implications for clinical enrollments for both AD and LBD-specific pathologies as well as clinical care. They also highlight the value of developing sensitive biofluid or imaging-based biomarkers for LBD.

## 2. Methods

### 2.1 Studied Participants

We focus on clinical case series at the Alzheimer’s Disease Centers (ADCs) from the National Alzheimer’s Coordinating Center (NACC), using longitudinal Uniform Data Set and the Neuropathology Data Set, collected up to August 20, 2019. This data includes records from 40,858 participants, 2,549 of which underwent brain autopsy post-mortem, and had AD and LBD pathologies evaluated based on brain histology^1^. We further limit our analysis to participants: (1) who had clinical evaluation and diagnosis for AD and/or LBD and (2) whose first record in the NACC data shows cognitively normal (CDR 0.0) or early dementia stages (CDR 0.5 or 1.0). This results in 1,887 participants in the final study.

### 2.2 Autopsy-Confirmed Outcome Definition

In our analysis, we define a participant outcome as AD, LBD, AD+LBD, and neither of them. We note that in this paper we do not focus on other dementia subtypes such as vascular dementia (VD) or frontotemporal lobar degeneration (FTLD). Definitions of each autopsy-confirmed subtype in our study are as follows.

**AD** - Participants has undergone autopsy and their NIA-AA Alzheimer’s disease neuropathologic change, ADNC-ABC score (Hyman et al. 2012) ^2^ is 2 (intermediate) or 3 (high), and their Lewy body pathology derived^3^ result is marked as 0 (No Lewy body pathology).

**LBD** - Participants has undergone autopsy and their NIA-AA Alzheimer’s disease neuropathologic change, ADNC-ABC score is either 0 (Not AD), or 1 (Low ADNC), and their Lewy body pathology derived results is marked as 3 (Neocortical - diffuse)

**AD + LBD** - Participants has undergone autopsy and their NIA-AA Alzheimer’s disease neuropathologic change (ADNC) (ABC score) is 2 (intermediate) or 3 (high); and their Lewy body pathology derived results is marked as 1 (Brainstem-predominan), 2 (Limbic - transitional) or 3 (Neocortical - diffuse).

**Neither** - Participants who have undergone autopsy and do not match any of the AD, LBD or AD+LBD criteria.

### 2.3 Clinical Diagnosis Identification

We define clinical diagnosis as either AD, LBD, AD+LBD, or neither. The clinical definitions are as follows:

**AD** - Clinician has marked the diagnosis as *Presumptive etiologic diagnosis of the cognitive disorder — Alzheimer’s disease*^4^; and the *Presumptive etiologic diagnosis of the cognitive disorder — Lewy body disease*^5^ is marked as 0 (No).

**LBD** - Clinician has marked the diagnosis as *Presumptive etiologic diagnosis of the cognitive disorder — Lewy body disease*^6^, and the *Presumptive etiologic diagnosis of the cognitive disorder — Alzheimer’s disease*^7^ is marked as 0 (No).

**AD + LBD** - Both positive diagnosis of *Presumptive etiologic diagnosis of the cognitive disorder — Alzheimer’s disease* and *Presumptive etiologic diagnosis of the cognitive disorder — Lewy body disease* are recorded.^8^

**Neither** - If none of the criteria above is met.

### 2.4 Evaluation Time Points

We evaluate the clinical diagnosis accuracy for the included participants at Cognitive Dementia Rating Global Score (Morris 1993) (CDR) of 0.5, 1.0, 2.0 and 3.0. At each cognitive impairment level, we evaluate clinical diagnosis accuracy at the first visit where the CDR score becomes equal to 0.5, 1.0, 2.0 or 3.0. We compare the clinical diagnosis against the autopsy results.

### 2.6 Diagnosis Disparities

We also measure clinical diagnosis disparities based on sexes, races, ages and education years. We report, per subgroup, the average metrics over all subtypes (if one subtype is missing, the average is computed based on other non-missing subtypes). Thus, the bootstrapping results show the distribution of average diagnosis performances. Besides metrics used in Table 3, we also compute False Negative Rate (FNR) for each group.

### 2.5 Statistical Analysis and Evaluation Metrics

All our results include confidence intervals, measured via bootstrapping method. Our bootstrapping algorithm includes 1000 iterations, and results are reported with 95% confidence intervals. P-values are computed based on one-sided testing and a 0.05 level indicates significance.

For evaluating accuracy of clinical diagnosis, we report positive predictive value, sensitivity, specificity and false negative rates, as well as confusion matrix between clinical diagnosis and autopsy results at various dementia stages (CDR 0.5, 1.0, 2.0 and 3.0). For those participants clinically diagnosed as *Neither* at different dementia stages, we further report the primary etiologic diagnosis^9^ made by the clinician instead.

## 3. Results

Overall, 1,887 participants met the criteria for inclusion, of which 757 had autopsy-confirmed AD, 46 had autopsy-confirmed LBD, 572 had autopsy-confirmed AD+LBD, and 512 were confirmed to have neither. Table 1 includes the characteristics for these participants. Participants who have LBD during autopsy are more likely to be male (74% vs 56% p-value: 0.007), and with fewer years of education (15.30 vs 16.42 p-value: 0.022). Participants with AD+LBD at autopsy have overall higher CDR at first visits (0.66 vs 0.59 p-value: <0.001).

**Table 1:** Participants Characteristics

| Characteristics | All Participants | Autopsy confirmed Alzheimer's Disease | Autopsy confirmed Lewy Body Disease | Autopsy confirmed AD+LBD | Neither AD nor LBD confirmed at autopsy |
| --- | --- | --- | --- | --- | --- |
| Total number of included participants | 1887 | 757 | 46 | 572 | 512 |

<sup>9</sup> NACCETPR in NACC data: https://files.alz.washington.edu/documentation/uds3-rdd.pdf

|  |  |  |  |  |  |
| --- | --- | --- | --- | --- | --- |
| **Sex** |  |  |  |  |  |
| Male | 1052(56%) | 384(51%) | 34(74%) | 326(57%) | 308(60%) |
| Female | 835(44%) | 373(49%) | 12(26%) | 246(43%) | 204(40%) |
| **Ethnicity** |  |  |  |  |  |
| White | 1765(94%) | 705(93%) | 44(96%) | 537(94%) | 479(94%) |
| Black or African American | 57(3.0%) | 24(3.2%) | 0(0.0%) | 18(3.2%) | 15(2.9%) |
| American Indian or Alaska Native | 2(0.11%) | 2(0.26%) | 0(0.0%) | 0(0.00%) | 0(0.00%) |
| Native Hawaiian or Pacific Islander | 1(0.05%) | 0(0.00%) | 0(0.0%) | 0(0.00%) | 1(0.20%) |
| Asian | 14(0.74%) | 4(0.53%) | 0(0.0%) | 5(0.87%) | 5(1.00%) |
| Multiracial | 35(1.85%) | 16(2.11%) | 2(4%) | 9(1.57%) | 8(1.56%) |
| Unknown or ambiguous | 13(0.69%) | 6(0.79%) | 0(0.0%) | 3(0.52%) | 4(0.78%) |
| **# of participants at each stage** |  |  |  |  |  |
| CDR 0.5 | 1243 | 507 | 35 | 326 | 375 |
| CDR 1.0 | 1342 | 563 | 26 | 461 | 292 |
| CDR 2.0 | 815 | 357 | 13 | 298 | 147 |
| CDR 3.0 | 601 | 258 | 8 | 212 | 123 |
| **Average age at initial visits** | 73.60 | 75.20 | 73.04 | 72.83 | 72.14 |
| **Average CDR at initial visits** | 0.59 | 0.59 | 0.52 | 0.66 | 0.51 |
| **Average education years** | 16.42 | 16.00 | 15.30 | 16.46 | 17.10 |

### 3.1 Evaluation of Clinical Diagnosis

Of 1,887 participants studied, 1,243 had a clinical visit at CDR 0.5; 1,342 participants had a clinical visit at CDR 1.0; 815 participants had a clinical visit at CDR 2.0; and 601 participants had a clinical visit at CDR 3.0.

We analyzed each dementia stage independently, and measured the accuracy of the clinical diagnosis for the first visit at each CDR-defined stage. Table 2 includes the distribution of clinical diagnosis results compared to the autopsy results. Table 3 includes measures of positive predictive value (PPV, Precision), sensitivity and specificity of the clinical diagnosis for each stage of dementia.

**Table 2:** Distribution of clinicians’ for CDR 0.5, 1.0, 2.0 and 3.0

|  |  | Clinical Diagnosis |  |  |  |  |
| --- | --- | --- | --- | --- | --- | --- |
| Autopsy Results |  | AD | LBD | AD+LBD | Neither | CDR |
|  | AD | 288 | 8 | 9 | 202 | 0.5 |
|  | LBD | 8 | 14 | 2 | 11 |  |
|  | AD+LBD | 200 | 26 | 9 | 91 |  |
|  | Neither | 114 | 15 | 4 | 242 |  |
|  | **AD** | 483 | 3 | 22 | 55 | 1.0 |
|  | **LBD** | 6 | 12 | 7 | 1 |  |
|  | **AD+LBD** | 371 | 33 | 27 | 30 |  |
|  | **Neither** | 112 | 18 | 5 | 157 |  |
|  | **AD** | 315 | 6 | 11 | 25 | 2.0 |
|  | **LBD** | 3 | 8 | 1 | 1 |  |
|  | **AD+LBD** | 252 | 14 | 18 | 14 |  |
|  | **Neither** | 49 | 16 | 4 | 78 |  |
|  | **AD** | 217 | 4 | 13 | 24 | 3.0 |
|  | **LBD** | 1 | 7 | 0 | 0 |  |
|  | **AD+LBD** | 172 | 19 | 10 | 11 |  |
|  | **Neither** | 38 | 5 | 3 | 77 |  |

**Table 3:**
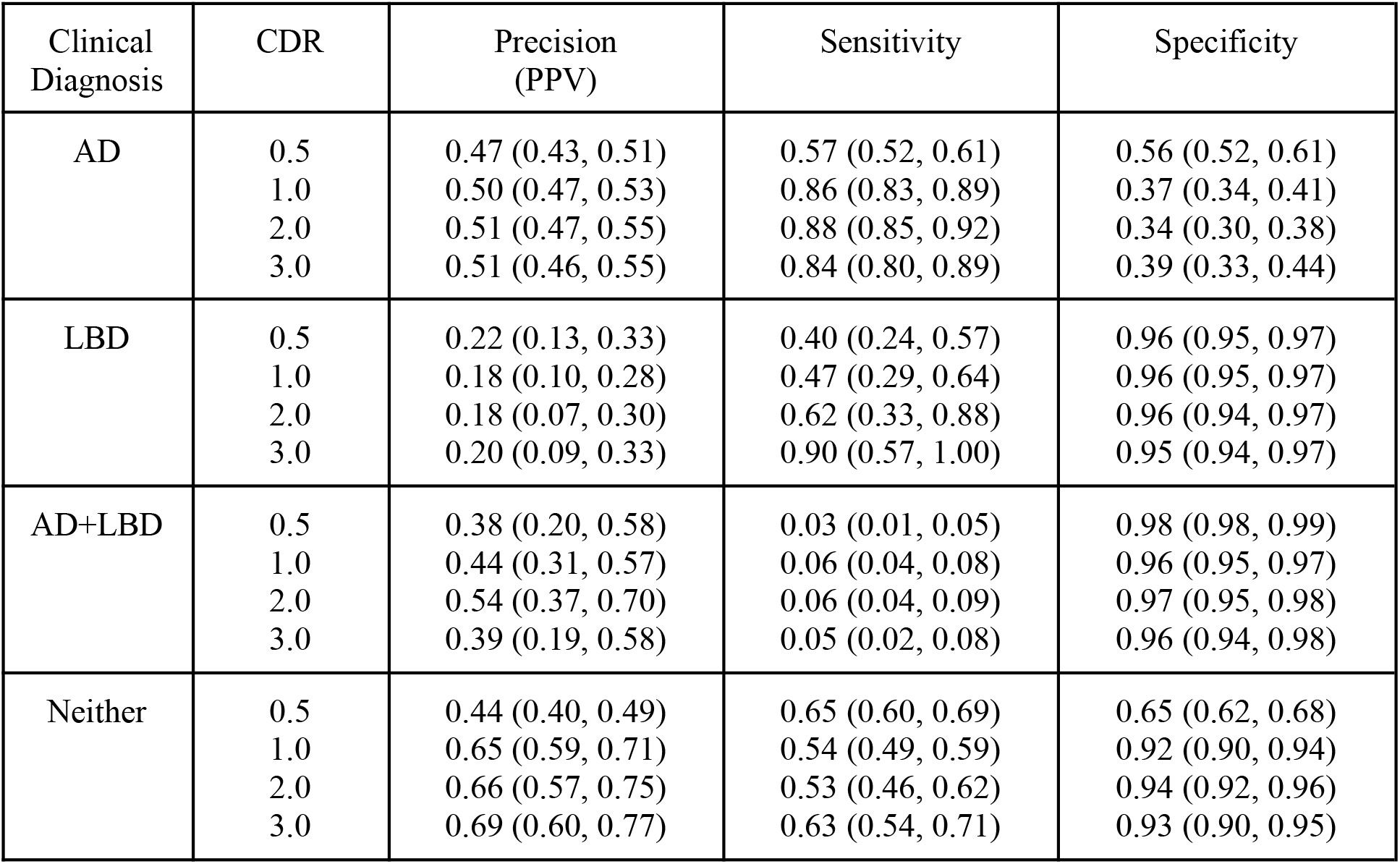
Clinical Diagnosis Performance for All Dementia Stages All Subtypes

| Clinical Diagnosis | CDR | Precision (PPV) | Sensitivity | Specificity |
| --- | --- | --- | --- | --- |
| AD | 0.5 | 0.47 (0.43, 0.51) | 0.57 (0.52, 0.61) | 0.56 (0.52, 0.61) |
|  | 1.0 | 0.50 (0.47, 0.53) | 0.86 (0.83, 0.89) | 0.37 (0.34, 0.41) |
|  | 2.0 | 0.51 (0.47, 0.55) | 0.88 (0.85, 0.92) | 0.34 (0.30, 0.38) |
|  | 3.0 | 0.51 (0.46, 0.55) | 0.84 (0.80, 0.89) | 0.39 (0.33, 0.44) |
| LBD | 0.5 | 0.22 (0.13, 0.33) | 0.40 (0.24, 0.57) | 0.96 (0.95, 0.97) |
|  | 1.0 | 0.18 (0.10, 0.28) | 0.47 (0.29, 0.64) | 0.96 (0.95, 0.97) |
|  | 2.0 | 0.18 (0.07, 0.30) | 0.62 (0.33, 0.88) | 0.96 (0.94, 0.97) |
|  | 3.0 | 0.20 (0.09, 0.33) | 0.90 (0.57, 1.00) | 0.95 (0.94, 0.97) |
| AD+LBD | 0.5 | 0.38 (0.20, 0.58) | 0.03 (0.01, 0.05) | 0.98 (0.98, 0.99) |
|  | 1.0 | 0.44 (0.31, 0.57) | 0.06 (0.04, 0.08) | 0.96 (0.95, 0.97) |
|  | 2.0 | 0.54 (0.37, 0.70) | 0.06 (0.04, 0.09) | 0.97 (0.95, 0.98) |
|  | 3.0 | 0.39 (0.19, 0.58) | 0.05 (0.02, 0.08) | 0.96 (0.94, 0.98) |
| Neither | 0.5 | 0.44 (0.40, 0.49) | 0.65 (0.60, 0.69) | 0.65 (0.62, 0.68) |
|  | 1.0 | 0.65 (0.59, 0.71) | 0.54 (0.49, 0.59) | 0.92 (0.90, 0.94) |
|  | 2.0 | 0.66 (0.57, 0.75) | 0.53 (0.46, 0.62) | 0.94 (0.92, 0.96) |
|  | 3.0 | 0.69 (0.60, 0.77) | 0.63 (0.54, 0.71) | 0.93 (0.90, 0.95) |

### 3.2 Clinically Missed Diagnosis

We further investigated the alternative clinical diagnosis made by the clinicians instead of the true underlying autopsy-confirmed AD, LBD or AD+LBD (i.e corresponding to the “Neither” column in Table 2). The histogram of clinically-primary etiologic diagnosis made for these cases are included in Figure 1. We report the results at different CDR stages, for three autopsy-confirmed subtypes (i.e. AD, LBD and AD+LBD). Of note, other than AD and/or LBD, the most common alternative clinical diagnoses missing pathological AD, LBD, and AD+LBD were behavioral variant frontotemporal dementia (bvFTD) or primary progressive aphasia (PPA).

**Figure 1:**
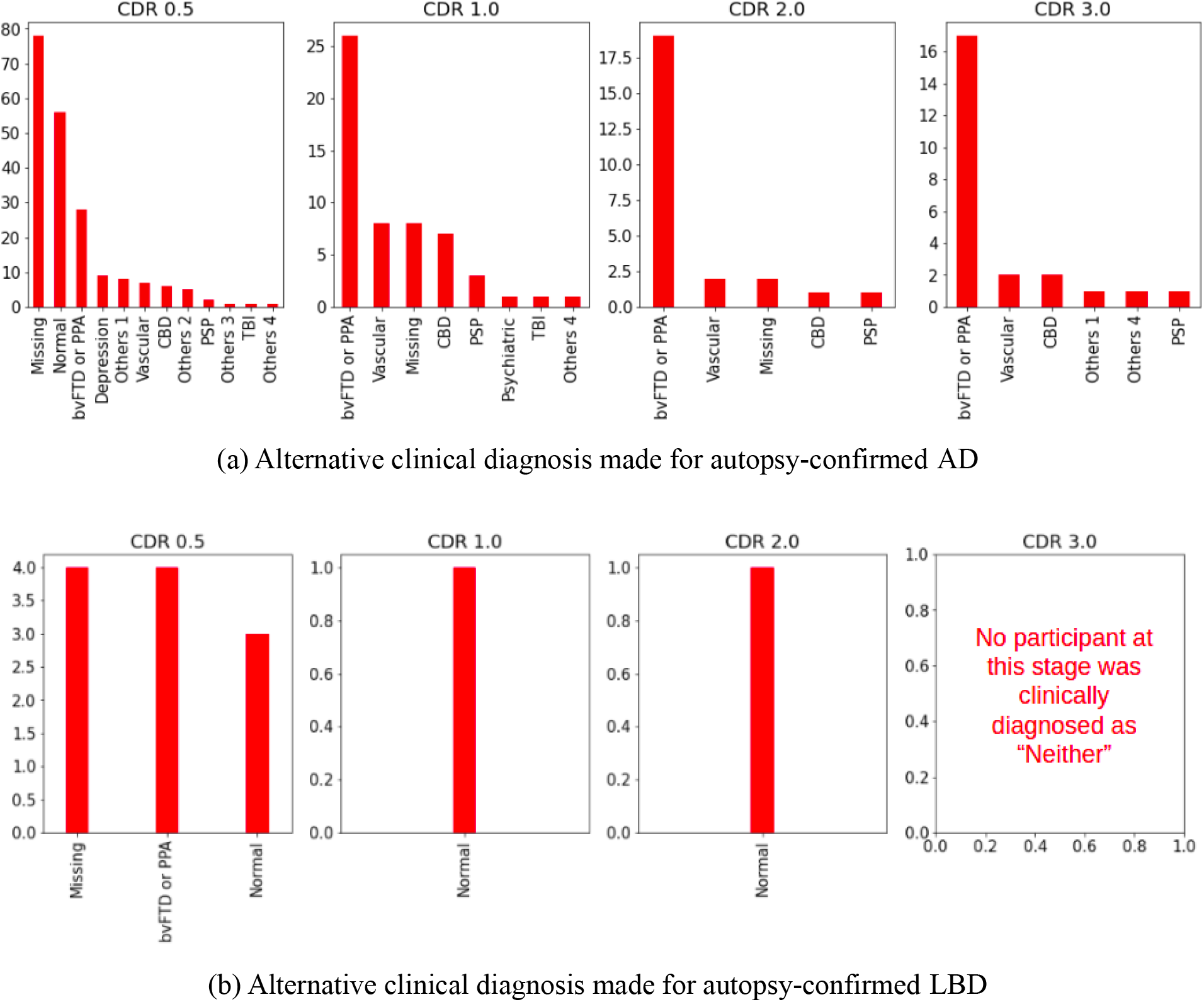
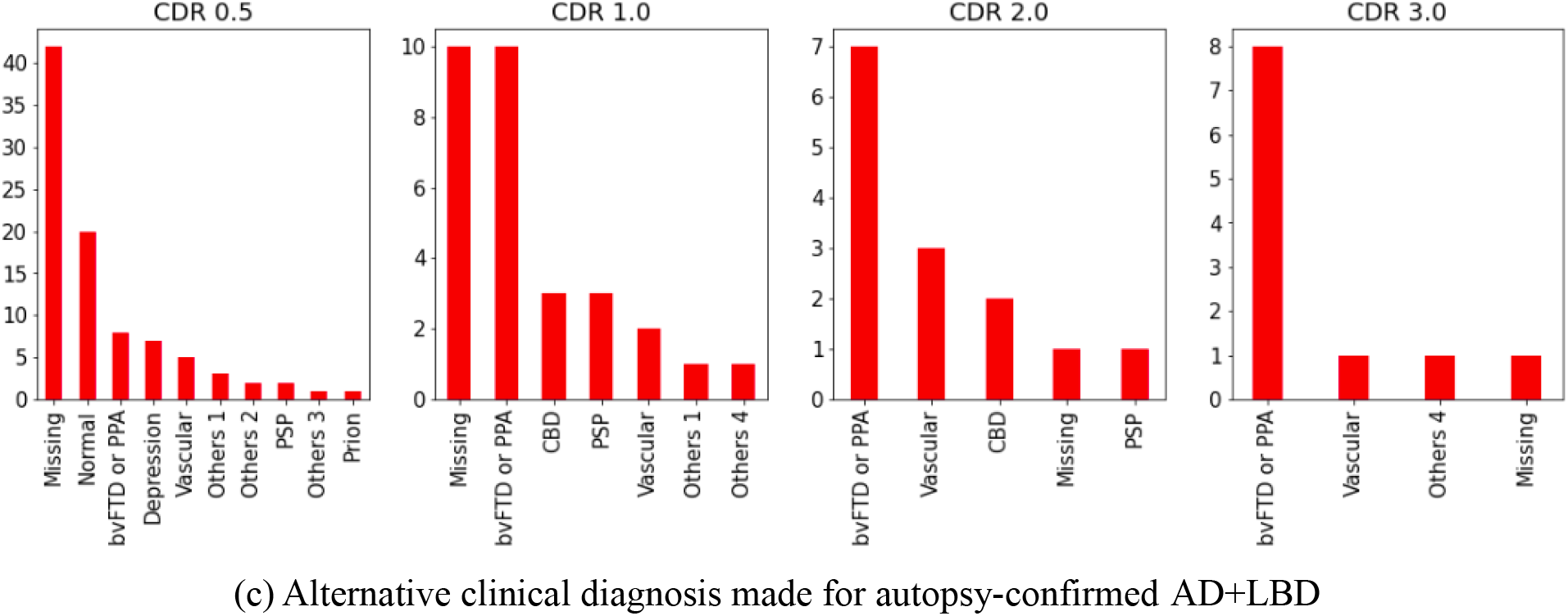
Distribution of clinically primary etiology diagnosis when pathological AD, LBD or AD+LBD was missed by clinicians. The results are analyzed for the first visits of each CDR stage of each subtype (i.e. AD, LBD, AD+LBD). For the corresponding clinically-primary etiology diagnosis names in NACC dataset, please refer to Table S1 in the Supplementary Document.

### 3.2 Diagnosis Disparities

Figure 2 shows the average false negative rate (FNR), sensitivity, positive predictive value (precision) and specificity across all four classes (AD, LBD, AD+LBD, Neither) for different subgroups based on sex, race, age, and education years. The average diagnosis quality (both in terms of sensitivity and false negative rate) significantly improves for males as dementia stage progresses, whereas this is not observed for female participants. Similarly, the diagnosis accuracy increases in White participants as the dementia progresses, but this trend is reversed in Blacks/African-Americans and other races/ethnicities. Black/African-American participants receive a clinical diagnosis that is less accurate compared to White participants, even at later dementia stages (median FNR, sensitivity, PPV and specificity disparity at CDR 2.0: 24.7%, 24.7%, 33.3%, 17.1%; CDR 3.0: 31.7%, 31.7%, 30.8%, 17.5%). Higher education levels do not improve clinical diagnosis accuracy, although we can see a trend of better sensitivity for participants with higher than master’s degree (education years: >16) at higher dementia stages. Also, although age does not have a significant impact on clinical diagnosis, the average diagnosis accuracy for older participants (>80) tends not to be as accurate as other age groups with regard to sensitivity and positive predictive values, especially at later stages.

**Figure 2:**
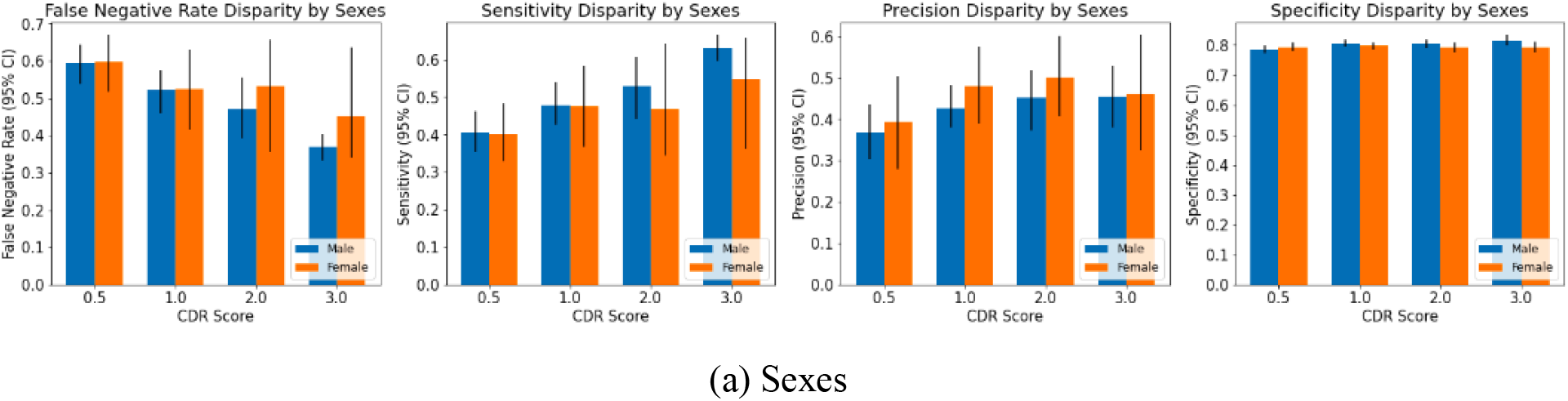
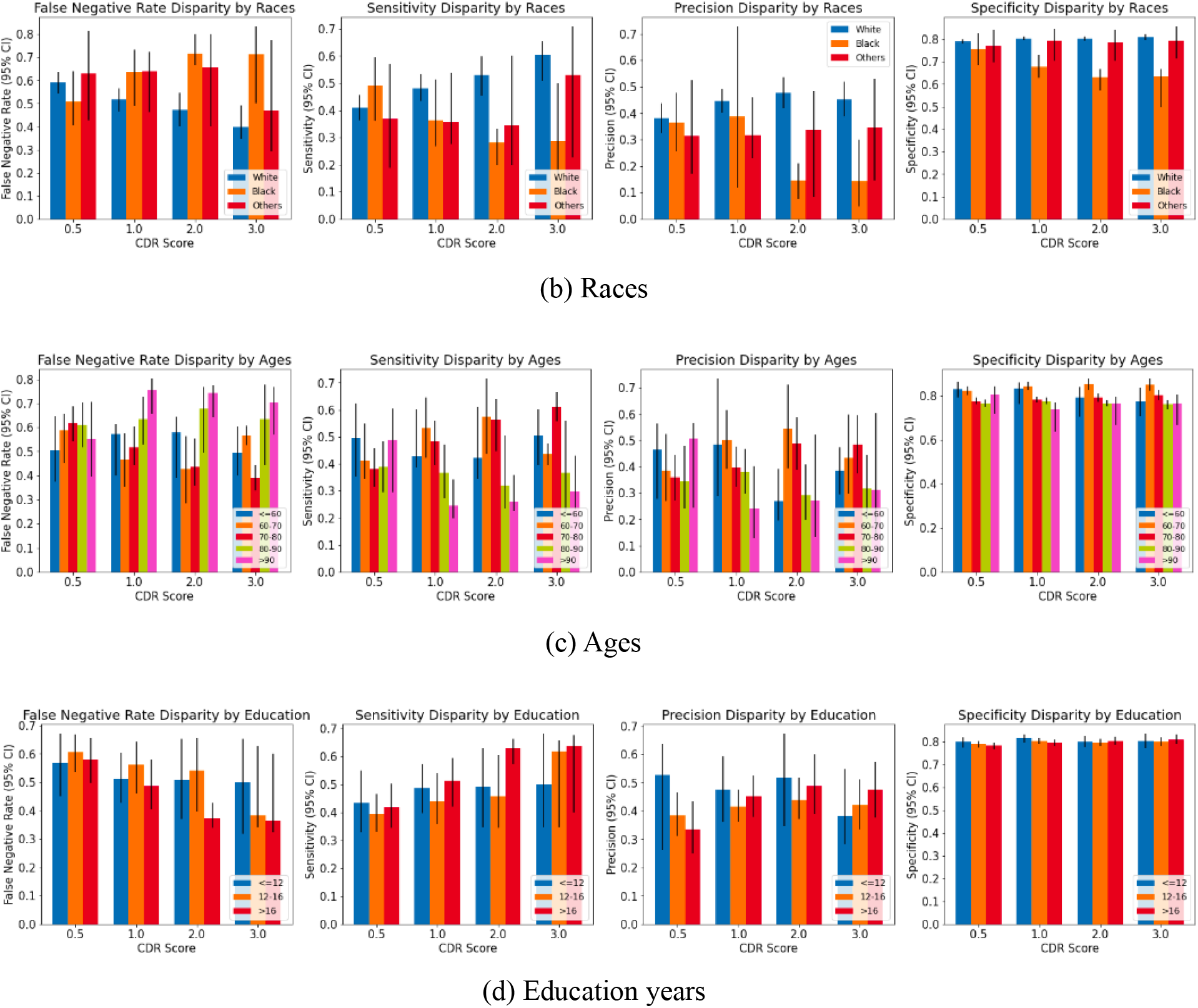
Clinician diagnosis disparities by (a) sexes, (b) races, (c) ages, and (d) education years. Disparities are evaluated by false negative rate, sensitivity, precision and specificity (from left to right at each row). Error bars indicate 95% confidence intervals.

## 4. Discussion

Dementia has remained an incurable disease despite massive burden to patients, their caregivers and to the society. Despite national investments, clinical trials to find a treatment have not made a breakthrough (Alexander, Emerson, and Kesselheim 2021) (Carroll 2017) (Liu et al. 2019). Our results indicate that clinical diagnosis has limitations in identifying distinct neuropathologies that are known markers for Alzheimers’ disease, Lewy body disease and the Lewy body variant of Alzhimer’s Disease. This has important implications for clinical trials for agents that target β-amyloid clearance (Ostrowitzki et al. 2017) (Egan et al. 2019) (Honig et al. 2018) (NIA 2021) (Huang, Chao, and Hu 2020) (Kurz and Perneczky 2011) or other subtype-specific neuropathologies. Indeed, we find that of participants diagnosed with AD in the clinic, 33% (CDR 0.5) to 41% (CDR 2.0) had AD+LBD pathology. In the absence of PET imaging tracers for Lewy bodies, using β-amyloid or Tau tracers with PET imaging to recruit Alzheimer’s disease participants (Klunk et al. 2004) can not identify these with AD+LBD. At the moment, most participants with AD+LBD autopsy were diagnosed as AD (from 61% at CDR 0.5 to 85% at CDR 2.0). Positive predictive values for clinical diagnosis of AD+LBD were low at different dementia stages, ranging from 37.5% to 53.5%. Sensitivity of clinical diagnosis for AD+LBD was poor, ranging from 2.7% (95% CI: 1.2%, 4.8%) at CDR 0.5, to 4.7% (95% CI: 2.1%, 7.8%) at CDR 3.0. And while autopsy-confirmed AD+LBD was more common than LBD (572 vs 46 overall), this prevalence did not translate to the clinic. The inaccuracies of clinical diagnosis for AD+LBD have also been recognized in previous smaller studies (Selvackadunco et al. 2019) although regardless of dementia stages. Our finding on much larger data shows that these inaccuracies do not improve as participants’ impairment increases.

Besides the challenges revealed in clinical diagnosis of AD+LBD, we also find that even Alzheimer’s disease diagnosis is difficult. At early dementia stages (CDR 0.5), clinical diagnosis of AD had low sensitivities 56.8% (95% CI: 52.4%, 61.1%) and specificities 56.2% (95% CI: 52.4%, 61.1%). While sensitivity improved at more advanced dementia stages (CDR 3.0), specificity further dropped to 38.5% (95% CI: 33.4%, 43.6%). When analysing alternative clinical diagnosis made instead of AD, we see “Normal” diagnosis (i.e. no impairment); primary progressive aphasia (PPA) or behavioral variant frontotemporal dementia (bvFTD) were the leading primary etiologic diagnosis when the neuropathological AD was missed. While sensitivity and specificity are important clinically, for clinical trial enrollments, positive predictive values are more important. PPV of clinical diagnosis of AD was only 47.2% (95% CI: 43.3%, 51.2%) at CDR 0.5, and even at higher dementia stages (CDR 3.0), this accuracy remained low at 50.8% (95% CI: 45.9%, 55.4%). This means that of every two participants enrolled in an AD-specific clinical trial, one has neuropathologies other than AD, and will likely not experience benefits from the AD-specific treatments.

Lewy body disease was more commonly manifested as AD+LBD (46 vs 572 Table 1), which as we saw, was often clinically diagnosed as AD only. However, focusing on participants with clinically diagnosed LBD alone without AD diagnosis, we observe overall poor positive predictive values of only 22.3% (95% CI: 12.5%, 33.3%) to 19.5% (95% CI: 8.6%, 33.3%) at dementia stages CDR 0.5 to 3 respectively. Of these participants, majority (32% at CDR 2.0 to 54% at CDR 3.0) harboured AD pathology, which was also indicated by (Selvackadunco et al. 2019). This may negatively impact clinical trials that enroll LBD participants at early stages. (Goldman et al. 2020) (Lee et al. 2019).

A trend similarly observed for AD, LBD and AD+LBD is that when missed by the clinicians the most common alternative diagnosis after “No impairment” or “Undecided” is the Behavioral variant FTD syndrome (bvFTD) or Primary progressive aphasia (PPA). While frontotemporal dementia is not the focus of our current study, our finding indicates potential over-diagnosis of this dementia variant at all levels of impairment.

Finally, we highlight the results of the analysis on disparities of clinical diagnosis accuracy according to sex, race, age, and education years. Our results confirm previous studies (Alzheimer’s Association 2021) (Lin et al. 2020) (Gianattasio et al. 2019) (Husaini et al. 2003) by showing that for minorities (Women, Black, Older and Lower educated participants) the current clinical diagnostic is not sufficiently accurate at various stages of impairment. Improving the diagnostic instruments such as MMSE (Chin, Negash, and Hamilton 2011) (Wood et al. 2006) (Manly and Espino 2004) (Pedraza et al. 2012), CDR, and developing more accurate imaging and cerebrospinal fluid biomarkers particularly for minority populations will be important steps towards better screening, diagnosis and treatment design for dementia.

Our study has several limitations: First, the cohort in our study is relatively small, primarily white and does not reflect the real-world population distribution. Our disparity analysis suggests that the clinical diagnostic accuracies for minority populations are lower than other subgroups, which further suggests that the accuracies of clinical diagnosis reflecting real-world ethnicity distributions are even lower than reported here. Second, when analyzing the diagnosis disparity for different groups, we only report the average metric over four subtypes (AD, LBD, AD+LBD, Neither). Further breakdown of each subtype was not possible due to the small sample size in our study. However, we acknowledge that the metrics reported likely will be different for different dementia subtypes. Third, other dementia subtypes (particularly vascular dementia and Frontotemporal dementia) are also important and necessary to study, however we only focused on AD, LBD, and AD+LBD in this study. We will expand similar analysis to other subtypes in future studies. Lastly, the dataset we used was focusing on AD research. In the clinic, diagnostic accuracy may be worse given less clinical information and less expertise of practitioners. Furthermore, the heterogeneity of cases will likely be higher in a clinic based population. A research cohort may be biased to healthier participants less likely to have comorbidities that would further mask diagnostic accuracy.

In conclusion, our study reveals important shortcomings pertaining to accurate and unbiased clinical diagnosis for Alzheimer’s disease and Lewy body disease at all dementia stages. Our results motivate use of machine learning and AI in improving the clinical diagnosis of dementia subtypes.

## Data Availability

All the data are available by requesting from NACC

https://naccdata.org/

## Acknowledgments

Study members HW and NR were fully and partially supported by Leon Lowenstein Foundation during the course of this study. NR and AVM were also partially supported by NIH/NIA P30AG066512. The NACC database is funded by NIA/NIH Grant U01 AG016976. NACC data are contributed by the NIA-funded ADRCs: P30 AG019610 (PI Eric Reiman, MD), P30 AG013846 (PI Neil Kowall, MD), P50 AG008702 (PI Scott Small, MD), P50 AG025688 (PI Allan Levey, MD, PhD), P50 AG047266 (PI Todd Golde, MD, PhD), P30 AG010133 (PI Andrew Saykin, PsyD), P50 AG005146 (PI Marilyn Albert, PhD), P50 AG005134 (PI Bradley Hyman, MD, PhD), P50 AG016574 (PI Ronald Petersen, MD, PhD), P50 AG005138 (PI Mary Sano, PhD), P30 AG008051 (PI Thomas Wisniewski, MD), P30 AG013854 (PI Robert Vassar, PhD), P30 AG008017 (PI Jeffrey Kaye, MD), P30 AG010161 (PI David Bennett, MD), P50 AG047366 (PI Victor Henderson, MD, MS), P30 AG010129 (PI Charles DeCarli, MD), P50 AG016573 (PI Frank LaFerla, PhD), P50 AG005131 (PI James Brewer, MD, PhD), P50 AG023501 (PI Bruce Miller, MD), P30 AG035982 (PI Russell Swerdlow, MD), P30 AG028383 (PI Linda Van Eldik, PhD), P30 AG053760 (PI Henry Paulson, MD, PhD), P30 AG010124 (PI John Trojanowski, MD, PhD), P50 AG005133 (PI Oscar Lopez, MD), P50 AG005142 (PI Helena Chui, MD), P30 AG012300 (PI Roger Rosenberg, MD), P30 AG049638 (PI Suzanne Craft, PhD), P50 AG005136 (PI Thomas Grabowski, MD), P50 AG033514 (PI Sanjay Asthana, MD, FRCP), P50 AG005681 (PI John Morris, MD), P50 AG047270 (PI Stephen Strittmatter, MD, PhD).

## Footnotes

1 Neuropathology data was limited to participants who had completed NACC Neuropathology Form version 10.

2 NPADNC in NACC Neuropathology data: https://files.alz.washington.edu/documentation/rdd-np.pdf

3 NACCLEWY in NACC Neuropathology data: https://files.alz.washington.edu/documentation/rdd-np.pdf

4 NACCALZD=1 in NACC data: https://files.alz.washington.edu/documentation/uds3-rdd.pdf

5 NACCLBDE=0 or 8 in NACC data: https://files.alz.washington.edu/documentation/uds3-rdd.pdf

6 NACCLBDE=1 in NACC data: https://www.alz.washington.edu/WEB/rdd_uds.pdf

7 NACCALZD=0 or 8 in NACC data: https://www.alz.washington.edu/WEB/rdd_uds.pdf

8 NACCALZD=1 and NACCLBDE=1 in NACC data: https://www.alz.washington.edu/WEB/rdd_uds.pdf

9 NACCETPR in NACC data: https://files.alz.washington.edu/documentation/uds3-rdd.pdf

