## Supplemental Table S1 for "On Gaps of Clinical Diagnosis of Dementia Subtypes: A Study of Alzheimer’s Disease and Lewy Body Disease"

**Supplementary Material**

Table S1: Primary etiologic diagnosis names[1] in the NACC dataset for Figure 1

| Name in Figure 1 | Name in the NACC dataset |
|---|---|
| Missing | Missing/unknown |
| Normal | Not applicable, not cognitively impaired |
| bvFTD or PPA | FTLD, other |
| Depression | Depression |
| Vascular | Vascular brain injury or vascular dementia including stroke |
| CBD | Corticobasal degeneration (CBD) |
| PSP | Progressive supranuclear palsy (PSP) |
| TBI | Traumatic brain injury (TBI) |
| Psychiatric | Other psychiatric disease |
| Prion | Prion disease (CJD, other) |
| Others 1 | Cognitive impairment for other specified reasons (i.e., written-in values) |
| Others 2 | Cognitive impairment due to systemic disease or medical illness |
| Others 3 | Cognitive impairment due to medications |
| Others 4 | Other neurologic, genetic, or infectious condition |

[1] NACCETPR in NACC UDS-RDD form: https://files.alz.washington.edu/documentation/uds3-rdd.pdf
